# Cross-platform Clinical Proteomics using the Charité Open Standard for Plasma Proteomics (OSPP)

**DOI:** 10.1101/2024.05.10.24307167

**Authors:** Ziyue Wang, Vadim Farztdinov, Ludwig Roman Sinn, Pinkus Tober-Lau, Daniela Ludwig, Anja Freiwald, Fatma Amari, Kathrin Textoris-Taube, Agathe Niewienda, Anna Sophie Welter, Alan An Jung Wei, Luise Luckau, Florian Kurth, Matthias Selbach, Johannes Hartl, Michael Mülleder, Markus Ralser

## Abstract

The role of plasma and serum proteomics in characterizing human disease, identifying biomarkers, and advancing diagnostic technologies is rapidly increasing. However, there is an ongoing need to improve proteomic workflows in terms of accuracy, reproducibility, platform transferability, and cost-effectiveness. Here, we present the Charité *<u>O</u>pen Peptide <u>S</u>tandard for <u>P</u>lasma <u>P</u>roteomics* (OSPP), a panel of 211 extensively pre-selected, stable-isotope-labeled peptides combined in an open, versatile, and cost-effective internal standard for targeted and untargeted proteomic studies. The selected peptides are well suited for chemical synthesis, and distribute well over the captured analytical dynamic range and chromatographic gradients, and show consistent quantification properties across platforms, in serum, as well as in EDTA-, citrate, and heparin plasma. Quantifying proteins that function in a wide range of biological processes, including several that are routinely used in clinical tests or are targets of FDA-approved drugs, the OSPP quantifies proteins that are important for human disease. On an acute COVID-19 in-patient cohort, we demonstrate the application of the OSPP to i) achieve patient classification and biomarker identification ii) generate comparable quantitative proteomics data with both targeted and untargeted approaches, and iii) estimate peptide quantities for successful cross-platform alignment of proteomic data. The OSPP adds low costs per proteome sample, thus making the use of an internal standard accessible. In addition to the standards, corresponding spectral libraries and optimized acquisition methods for several platforms are made openly available.

## Introduction

The analysis of human plasma and serum proteomes is of increasing importance for biomedical research due to the minimally invasive sample collection of plasma and serum combined with the properties of the plasma proteome to directly reflect human physiological and/or pathophysiological states ^1–4^. Plasma proteomes reflect various physiological processes and plasma proteins can function as biomarkers for different conditions. This includes well-established biomarkers such as troponin T, C-reactive protein, procalcitonin, and cystatin C, supporting decision-making related to diagnosis, prognosis, and intervention monitoring ^5–9^.

Liquid chromatography-mass spectrometry (LC-MS) allows the simultaneous evaluation of large numbers of proteins, elucidating the functional implications of specific proteins, and unveiling changes within pertinent pathways while maintaining relatively low operational costs ^3,10–12^. Nevertheless, the routine implementation of LC-MS, especially discovery proteomics, still faces challenges arising from technical variances in acquisition batches and methods, analytical platforms, and processing pipelines. The presence of these variations results in incomparable relative quantification of proteins across platforms, adding complications to the data analysis and the routine implementation of the proteomic workflows.

A strategy for addressing these challenges is to aim for absolute rather than relative quantification methods. In mass spectrometry, isotope-labeled internal standards have been in use since the 1990s and allow the normalization of endogenous signals with absolute reference values ^13,14^. In proteomics, the spike in isotope-labelled peptide standards (SIS) where ^13^C, and ^15^N are commonly used stable isotopes ^15–19^ are used as quality control procedures for estimating absolute quantities at the peptide level. These help to achieve cross-platform and cross-laboratory reproducibility^16,20–22^. Commercially available peptide standards, such as the PQ500 standard (Biognosys, Switzerland) ^23–25^ or the PeptiQuant 270-protein human plasma MRM panel (MRM Proteomics Inc., Canada) ^26,27^, are designed for targeted analysis of broad sets of proteins. However, there is a continuing need to reduce the costs per sample, to improve the cross-platform detectability of the standards, and to improve data analysis strategies to efficiently make use of internal standards not only in targeted but also in discovery proteomics.

In response to these challenges, we introduce the Charité *<u>O</u>pen Peptide <u>S</u>tandard for <u>P</u>lasma <u>P</u>roteomics* (OSPP). Consisting of 211 isotope-labelled peptides derived from proteins involved in a wide range of biological processes. These peptides are selected to exhibit high analytical performance across various analytical platforms and blood matrices. Additionally, they are selected to be easily and economically synthesized, have biological meaning, and are broadly applicable across various clinical contexts, allowing constant detection at low technical variance.

## Results

### Selection of Analytically Robust Peptide Standards from Large-Scale Proteomic Data Sets

To assemble an efficient peptide standard for neat plasma proteomics, we utilized proteomic data produced in 1,505 control injections, which were measured alongside the analysis of 15,617 plasma and serum proteomes at the Charité High-throughput Mass Spectrometry Core Facility from 2020 to 2022. These samples were pools from prepared from human plasma and serum using a semi-automated workflow in 96-well plates which involves a clean-up step using solid phase extraction before being analyzed on a proteomic platform that uses analytical flow rate reverse phase chromatography with water-to-acetonitrile gradients and a throughput of 3-5 minutes/sample ^28^. Proteomes were recorded using data-independent acquisition on two SCIEX TripleTOF 6600+ instruments operating in SWATH ^29^ or Scanning SWATH ^28^ mode. Data was analyzed using DIA-NN ^30^ with the DiOGenes spectral library ^31^.

To prioritize the most reliably quantified precursors, we introduced a relative rank metric. This metric was calculated by first defining the precursor weight as the ratio of the precursor’s percentage presence (%*PPres*) to its coefficient of variation (%CV).

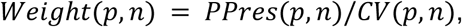

and a weight-based rank *Rank*(*p*, *n*) = *rank*{*Weight*(*p*, *n*)}. Here, *p* stands for precursor and *n* for a study pool series. The weight thus corresponds to a precursor’s signal-to-noise ratio (*S*/*N* = 1/*CV*) multiplied by its presence.

To exclude the influence of the total number of precursors on the ranking, we introduced relative rank *RelRank*(*p*, *n*), defined as the ratio of the rank to its maximum value in a study.

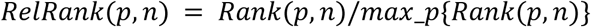

Finally, the precursor’s average (over considered studies) relative rank *RelRank*(*p*) was used to select the best non-project specific precursors for every protein while we also required that the lower cutoff of the relative rank be set as 0.6.

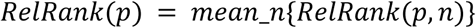

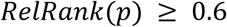

We selected only proteotypic peptides quantified in at least half of the examined studies. To allow coverage of a larger protein concentration range, only three top-ranked precursors were selected for each protein (Figure 1). Eventually, this selection process identified 382 consistently quantified peptides. We then ranked these according to their suitability as internal standards, such as a peptide length between 6-25 amino acids, a minimal likelihood of missed cleavages, and low susceptibility to chemical modifications (Figure 1, Supplementary Table 1). Among these, we further prioritized peptides with high synthesis efficiency according to the Peptide Analyzing Tool (Thermo Scientific) ^32^. We also incorporated 24 out of 50 peptides from a previous peptide panel, which showed excellent cross-platform analytical performances ^33,34^.

**Figure 1.**
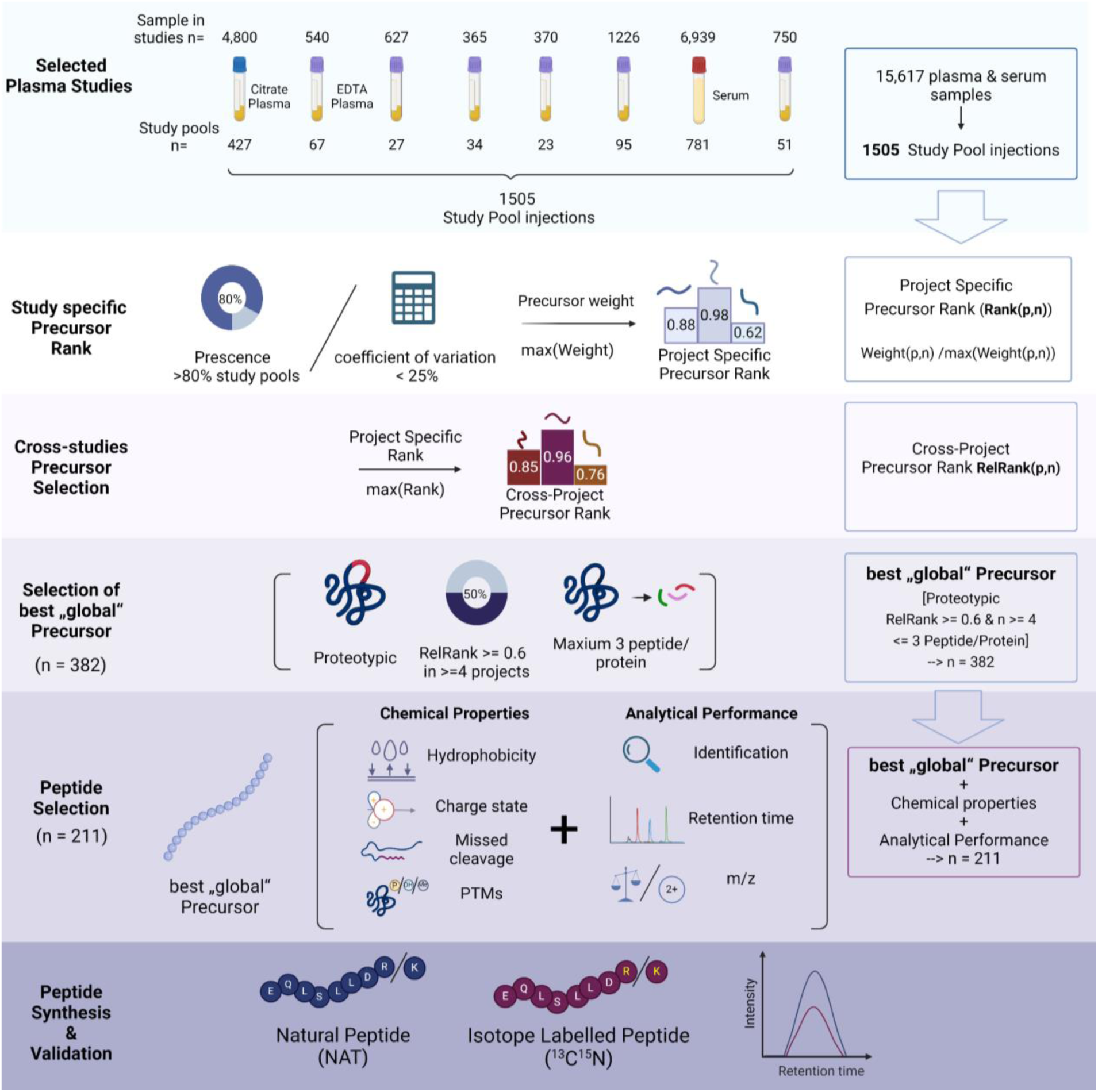
Overview of the Peptide Selection Process to generate the Charite Open Standard for Plasma Proteomics (OSPP) Plasma and serum proteomic data acquired from 1505 measurements of study control samples were used for selecting 187 peptides with ideal properties of an internal standard (Scheme). Peptide selection for the OSPP was based on consistent detection across studies, the obtained signal stabilities, chemical and biophysical properties, and suitability for synthesis. In addition, we added 24 peptides out of a previously generated targeted panel assay, which showed excellent cross-platform performance ^33^ The total of 211 selected peptides (Supplementary Table 1) were synthesized in both native and isotopically labeled form, which coeluted on the chromatogram.

Eventually, our selection converged on 211 proteotypic peptides (187 newly selected peptides plus 24 peptides which were already included in the previously described COVID-19/MPox panel ^33,34^. These peptides are derived from 131 plasma proteins, of which 57 key proteins are represented by two or three peptides. Due to the addition of the 24 best performing peptides from the COVID-19/MPox panel assay, for three proteins—SERPINA3, APOA1, and APOB— four or five peptides are included (Supplementary Table 1). The proteins cover four orders of magnitude in the concentration range of human plasma (Figure 2a) and have been associated with several diseases. They encompass enzymes, transporters, and cytokines (Figure 2b). They cover a broad range of FDA-approved drug targets ^35^ and a fraction already serve as routine clinical-chemistry biomarkers in different matrices (serum, citrate-, heparin-, or EDTA-plasma) (Figure 2b, lower panel).

**Figure 2.**
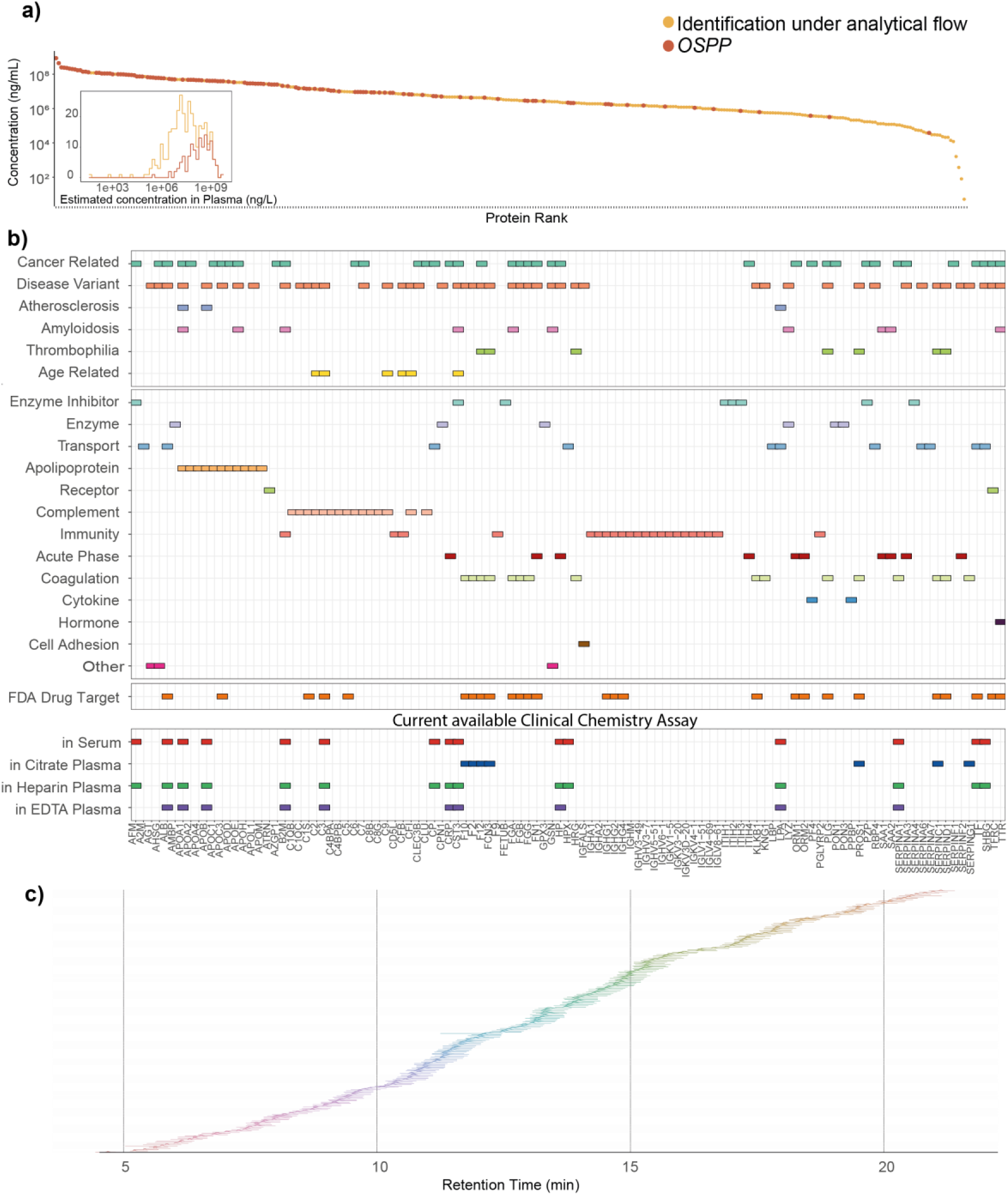
Characteristics of the OSPP peptide panel. **a)** Plasma proteins are represented by peptides in the OSPP, sorted by their abundance in human plasma according to analytical-flow rate DIA analysis (x-axis) as well as their estimated concentration on an absolute scale ^35,37^ (left y-axis). The inset shows the log-scale plasma protein concentration distribution of OSPP compared to those identified in the analytical flow DIA dataset. The panel peptides are derived from proteins whose concentration spans over four orders of magnitude. **b)** OSPP proteins and their associated biological functions as curated from The Human Protein Atlas ^35,37^ (upper panel), whether they are FDA-approved drug targets (middle panel), as well as their use in routinely used clinical tests based on Serum, Citrate, Heparin, or EDTA plasma, respectively (lower panel). **c)** Extracted ion chromatograms illustrating the chromatographic elution of the OSPP peptides in a 20-min µflow reversed-phase liquid chromatography, as analyzed by Zeno MRM-HR or Zeno SWATH DIA on a ZenoTOF 7600 instrument (SCIEX). Intensities were normalized to the maximum intensity of the respective peptide. The OSPP peptides distribute chromatographically.

### Assessment of OSPP Peptide Standards

To create the peptide panel, the selected peptides were synthesized in both native and isotopically labeled forms, with heavy labeling of the C-terminal arginine or lysine using ^13^C^15^N. Validation of the synthesized peptides involved quality checks via LC-UV/VIS by the peptide manufacturer and in-house analytical assessment by LC-MS analysis. Subsequently, native and SIS peptides were prepared in groups of 11 based on their abundance in EDTA plasma and analyzed with a 5 µl/min microflow-rate reversed-phase chromatographic gradient by ZenoSWATH-DIA on a ZenoTOF 7600 instrument (SCIEX) ^36^. Quality criteria entailed the co-elution with their respective native forms and absence of signals from native peptides. Reassuringly, the selected peptides all have well-distributed hydrophobicity scales, achieving a balanced elution distribution across the entire retention time range in a 20-minute active gradient microflow chromatography (Figure 2c, Supplementary Table 1).

Next we validated their performance in combination different anticoagulant matrices in human plasma. We evaluated the identification rate of the OSPP SIS upon spiking them into different blood-derived sample types matrices, Heparin-, Citrate-, and EDTA-plasma, a commercial plasma matrix (zenbio) as well as in serum. As an initial analysis of all stable isotopic labeled peptides, we added an equal concentration of each peptide (“Single-conc. Standard”, 21.1 ng total peptide amount) into 1.5 µg of total protein digest from each matrix. Most (199/211) of the peptides are well quantified in hexuplicate injections of commercial plasma and 97% (204 out of 211) peptides were detected in all tested anti-coagulant matrices using analytical flow rate chromatography (500 µl/min, 5-minute gradient) with a timsTOF HT mass spectrometer and analyzed using diaPASEF ^38^, demonstrating that the selected peptides are suitable as an internal standard in all the commonly used anticoagulant matrices and serum, as well as suit high-throughput acquisition methods. For those peptides that are not continuously identified, the reason falls on their low abundance in non-diseased blood samples (i.e. peptides from proteins such as CRP); or their closeness to detection limit resulted in their poor identification.

After evaluating signal intensities of each precursor from each OSPP SIS peptide in the different anticoagulant matrices, we created the concentration-matched OSPP, in which each signal corresponding to a stable isotope-labeled peptide matches its endogenous counterparts in EDTA plasma within one order of magnitude in signal intensity. For this, we divided the peptides into four concentration bins of 10 pg/µl to 2 ng/µl for each peptide in 10% (v/v) Acetonitrile (Supplementary Table 2). Notably, next the improving the analytical performances and to reduce the risk of signal suppression, the concentration matching also lowers the cost of the OSPP peptide panel, due to lower total quantities necessary.

To confirm the OSPP performance on different platforms, triplicates of eight-point external calibration curves were created for the 211 peptides, with BSA digest as a surrogate matrix. The non-labelled peptide standard in all calibration curves ranges from 0.5 ng to 0.032 pg, injected triplicates in each acquisition method. The limits of quantification (lower and upper: LLOQ and ULOQ) were determined based on the accuracy of replicated injection on the same LC-MS platform, and most peptides show a large concentration dynamic in their identification in neat plasma, with LLOQ less than 1 pg and ULOQ more than 500 pg (Supplementary Table 3).

### Evaluation of the OSPP in a case study of the human host response to SARS-CoV-2 infection

Next, we validated the OSPP for its application in targeted proteomics. As a use case, we focused on citrate-plasma samples obtained from a small (n=45) but well-balanced cohort of COVID-19 patients. We choose this cohort as a test case, as it contains samples from healthy individuals to severely ill patients, which 1) has similar effective size that is consistent in different severity groups, allowing fair comparison of the concentration changes of each OSPP peptide; 2) the inclusion of samples from healthy individuals to severely ill patients allows a wider range of concentration changes of OSPP peptides and identification of peptides that are only abundant in either healthy or diseased samples, avoiding missing values due to low abundance. The cohort comprises healthy controls as well as individuals hospitalized between March 1 and 26, 2020 at Charité ^28,33,39,40^ exhibiting varying severities of COVID-19, classified using the WHO ordinal scale for clinical improvement ^41,42^. The WHO severity ranges from 0 (healthy control), 3 (mild disease, hospitalized due to COVID-19, but without need for supplemental oxygen therapy), to 7 (critically ill patients with invasive mechanical ventilation and other organ support therapy) (Supplementary Table 4). Upon sample preparation using the aforementioned semi-automated workflow, 1 µl (40.4 ng, total peptide amount over all 211 peptides) of the OSPP was spiked into 1.5 µg of total plasma digest. Since targeted acquisition is commonly used as the method when coupling with stable isotope-labeled peptides, the samples were first separated using 20-min 5 µl/min µflow chromatography on an ACQUITY UPLC M-Class system (Waters), coupled with a ZenoTOF 7600 system (SCIEX). Data was recorded using a targeted method (Zeno MRM-HR) and processed using Skyline ^43^, including manual inspection of peak integration.

To evaluate how well peptides could be detected and quantified across healthy individuals or patients with varying COVID-19 severity, we used the Zeno MRM-HR method and tested for their presence in samples in the aforementioned cohort. Peptides that were detected in more than two-thirds of samples were included for subsequent analysis, and this criterion was met by 202 of the 211 peptides. Among those peptides, when only using the endogenous quantified value, half (106/202) were responsive depending on severity of symptoms. With the application of the OSPP as internal standards, technical bias and noise were better controlled, leading to more precise and reliable results. This improved measurement precision revealed 34 additional peptides with significant changes when quantified with MRM-HR, while 2 peptides which were previously associated with disease severity lost the significant association—likely these were called as false positives with the less precise method. As a result, more than two-thirds of peptides (138 out of 202) demonstrated significant changes correlated with COVID-19 severity (Supplementary Table 5. Moreover, principal component analysis of all peptide relative quantities normalized by OSPP grouped patients based on the WHO score, with the first dimension explaining 31.3% of the variance originating from disease severity (Figure 3b), of which we highlight the top 15 up- and down-regulated peptides that change according to WHO grade (Figure 3b). These peptides correspond to several proteins important for the host response in COVID-19. For example, peptides derived from tetranectin (CLEC3B) that modulate the biological function and inflammatory process during infection are down-regulated in severe COVID-19, while complement factor C3-derived peptides increased with severity (Figure 3c,d). Additionally, other peptides exhibited distinct signals corresponding to specific disease escalations. For example, a peptide derived from the enzyme inhibitor ITIH3 is strongly associated with infection itself, as is a peptide from the acute phase protein AHSG (Figure 3c,d). Others, such as peptides from the kidney and inflammation marker CST3, changed drastically during critical COVID-19 cases, indicating the possibility of kidney dysfunction in severe infection ^6^ and peptides associated with the calcium-regulated, actin-modulating protein gelsolin (GSN) whose reduced abundances were associated with worse outcomes ^44^ (Figure 3c,d).

**Figure 3.**
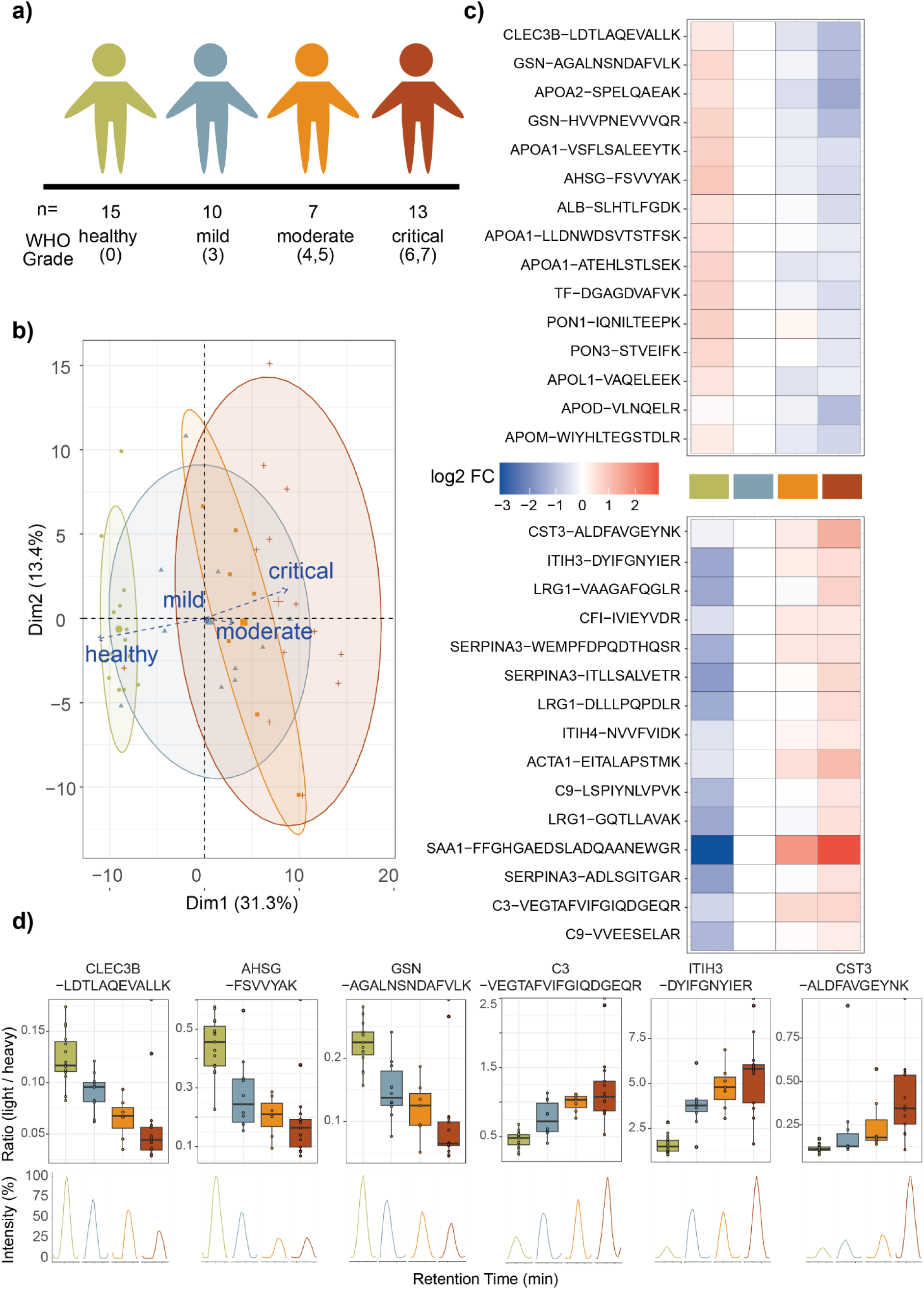
OSPP peptides measured by targeted proteomics report disease severity in a COVID-19 inpatient cohort. **a) Schematic illustration** Balanced cohort consisting of healthy volunteers and hospitalized patients suffering from acute COVID19 (a subgroup of the PA-COVID19 study ^39^) **b)** Unsupervised clustering by principal component analysis (PCA) based on the OSPP normalized quantity of 202 quantified peptides clusters patients with COVID-19 by disease severity. **c)** Peptides with a significant abundance change (down-(top panel) and up-regulated (bottom panel)) distinguish healthy from affected individuals, as well as mild from severe forms of the disease, represented by the WHO treatment escalation score. Heatmaps display the log2 fold-change of the indicated peptide to its median concentration in patients with a severity score of WHO3. The top 15 significant peptides (adjusted p < 0.05) are shown. **d)** Visualization of the response to COVID-19 based on selected peptides indicating different COVID-19 severity trends (changing with severity expressed according to the WHO ordinal scale (as in (c)), and differentiating healthy from COVID-19-infected individuals). Boxplots display the OSPP normalized quantity of selected peptides in patients in different severity groups, as explained in (a). The box-and-whisker plots display 25th, 50th (median), and 75th percentiles in boxes; whiskers display upper/lower limits of data (excluding outliers). The extracted ion chromatograms display the relative response of representative samples of individuals classified according to the disease severity.

In summary, OSPP could be effectively used in a targeted Zeno MRM-HR method to stratify the disease severity of COVID-19 patients. Furthermore, this targeted method successfully identified protein markers covered by OSPP for specific disease state transitions, as was reported in our previous discovery proteomics ^28,40^ and targeted MRM studies on the same cohort ^33^.

### Application of the OSPP for Cross-Method Proteome Analysis in Human plasma

To assess quantification performance across targeted and untargeted acquisition methods, we measured the same OSPP-spiked sample in both targeted (MRM-HR) and untargeted (ZenoSWATH-DIA ^36^) acquisition methods, first on the same acquisition platform (ZenoTOF 7600). For Zeno SWATH analysis, samples were acquired using the samples using the 20-minute 5 µl/min RP chromatographic gradient. The data processing pipeline involved DIA-NN ^30^, using a modification of the DiOGenes spectral library ^31^, to which we added isotopic labels (^13^C^15^N) specifically on OSPP peptides and excluded b-ions for quantification as they would have the same m/z for both SIS and native standards.

The majority of peptides (207/211), both the native and its matched SIS form, were quantified in over two-thirds of patient samples using Zeno SWATH DIA, with 199/211 of these pairs quantified consistently in both ZenoSWATH-DIA and Zeno MRM-HR, with the same fragment used for quantification (Figure 4a). Given that the source settings and ionization strategies were identical between the two acquisition methods, it is expected that shared fragments of each peptide would display a similar distribution between methods. Indeed, the overall fragment spectrum observed in ZenoSWATH-DIA closely matched that in Zeno MRM-HR (Figure 4b, Supplementary Table 6), with medians of relative signals from shared fragment ions displaying fold changes of each shared fragment in both acquisition methods (MRM-HR / ZenoSWATH-DIA) near 1 with both acquisition methods (0.9782 for endogenous and 0.9781 for heavy-labeled peptides). This similarity in fragment ion distributions confirms the consistency of OSPP’s performance between targeted and discovery methods and suggests low interference in DIA acquisition when using the OSPP standard, supporting its reliable technical performance in a discovery setup.

**Figure 4.**
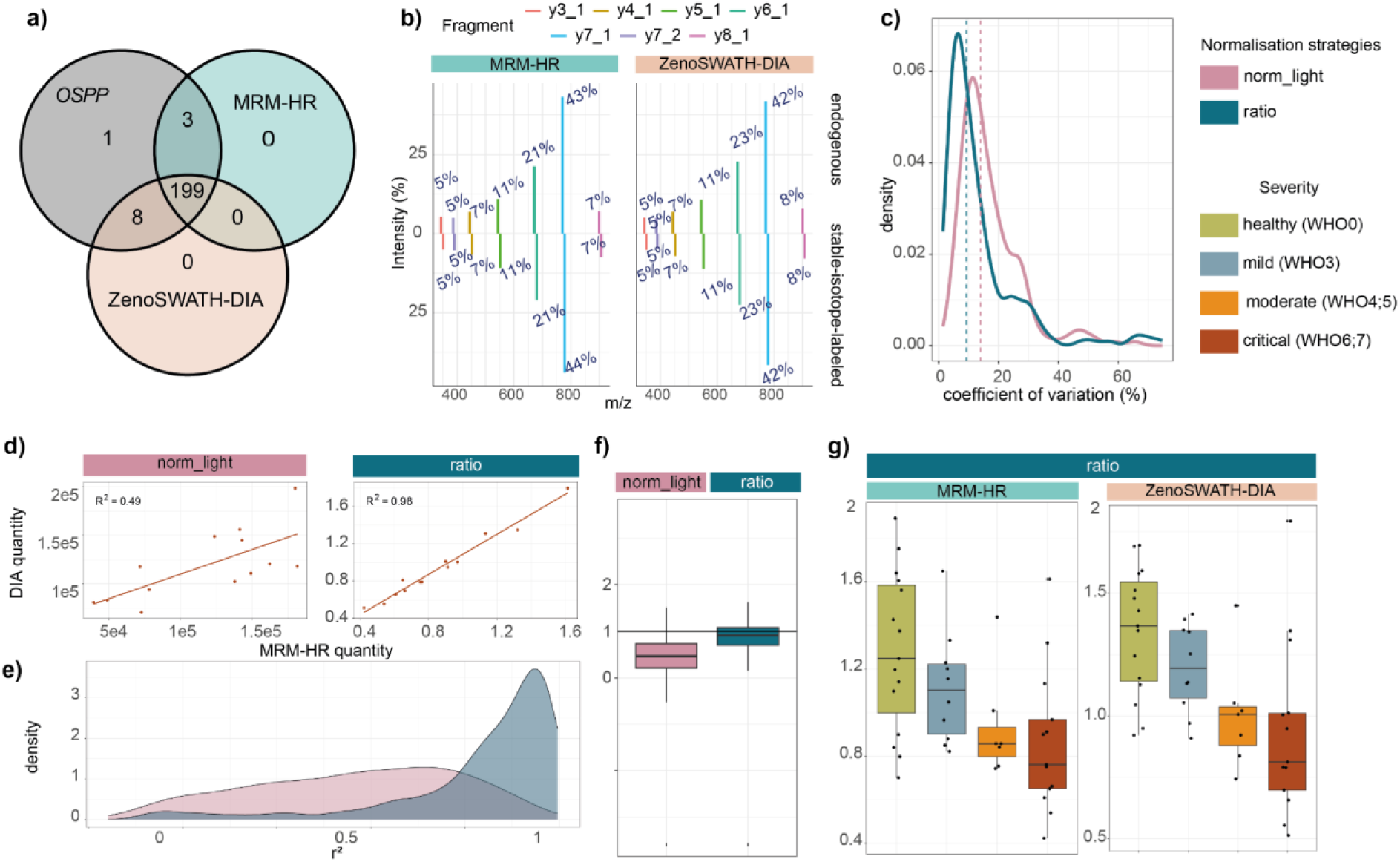
Using the OSPP as an internal standard to align targeted and untargeted MS data, on the example of a COVID-19 cohort. **a)** Intersections of quantified peptides (in both peptide groups, endogenous and SIS) of targeted and untargeted MS methods to the OSPP peptide list (Venn diagram) **b)** Average fragment spectral distribution of Alpha-2 macroglobulin-derived peptide FEVQVTVPK in triplicate study pool injection in both MRM-HR and ZenoSWATH-DIA. **c)** Coefficient of variation (CV) values calculated from each peptide quantity in quintuplicates of a study pool sample for both methods (density plot). Dashed lines display the median CVs of each normalization strategy (norm_light: 14%, ratio: 9%) **d)** Quantities of Alpha-2 macroglobulin-derived peptide FEVQVTVPK in critically ill COVID-19 patients (n = 13), as determined by Zeno MRM-HR and ZenoSWATH-DIA (correlation plot). A regression line was fitted to the data (y∼x). **e)** Distribution of all *r^2^* values calculated for each quantified light peptide in each severity group from correlating Zeno MRM-HR and ZenoSWATH-DIA (density plot, Supplementary Table 6). **f)** Distribution of slopes from correlating Zeno MRM-HR and ZenoSWATH-DIA, fitted to a linear model (y ∼ x) (Boxplot). The color of the dots represents the respective severity group. The box-and-whisker plots display 25th, 50th (median), and 75th percentiles in boxes; whiskers display upper/lower limits of data (excluding outliers). **g)** Median endogenous normalized (“norm_light”) and OSPP normalized (“ratio”) quantity of Alpha-2 macroglobulin-derived peptide FEVQVTVPK in patients across different severity groups (Boxplots). The box-and-whisker plots display 25th, 50th (median), and 75th percentiles in boxes; whiskers display upper/lower limits of data (excluding outliers).

To confirm that OSPP used in the DIA method also provides reliable peptide quantity results without notable interference, we first showcase the comparison of the quantities the DIA method acquired to those that are acquired in the targeted method. To confirm the usage of OSPP in supporting quantification consistency, we compared a normalization strategy that does not take into account the OSPP internal standard—median normalization via endogenous peptide quantities (referred to here as “norm_light”) to those that use OSPP as internal standard—ratio of the quantity of endogenous peptide over the quantity of its corresponding OSPP SIS (i.e., the light/SIS, “ratio”).

To assess the quantitative precision, we generated a study pool sample and analyzed it in quintuplicate, alongside the study samples. While ZenoSWATH-DIA produced precise quantification results also without the standard, a marked improvement in CV is observed upon normalizing the peptide signals to the OSPP. The median CV of all peptides quantified changed from 14% without the standard to 9% of the median CV with the OSPP normalized values (Figure 4c, Supplementary Table 6). Also, when comparing the fold change between peptide quantities in DIA and MRM-HR, the median fold change (DIA / MRM-HR) of the quantities for all OSPP-included peptides in all repeated injected samples is 0.006, indicating the inherit different quantification number between two acquisition methods, while after median normalization the median fold change increased to 1.18. When using OSPP as the internal standard for normalization, the median fold change of all peptides changed to closer to 1 (1.06) (Supplementary Table 6) which indicates a higher quantification consistency is reached with OSPP normalization. This result showed that OSPP used as an internal standard is compatible for DIA acquisition; it not only has low interference in DIA acquisition but also supports more consistent quantification between DIA and targeted methods.

Next, we explored quantitative similarities and differences between the targeted Zeno MRM-HR and the DIA method by fitting a linear regression model to each peptide quantity in each severity group. In our comparison, an *r^2^* close to 1 indicates data acquired from ZenoSWATH-DIA has a close relation to Zeno MRM-HR data, and a slope close to 1 indicates that the quantities obtained from both methods are highly consistent. For example, the quantity for the Alpha-2 macroglobulin (A2M)-derived peptide FEVQVTVPK correlated with an *r^2^*of 0.49 without OSPP normalization for Zeno MRM-HR and ZenoSWATH-DIA. When applying OSPP to form ratios between the endogenous peptide signal and the standard, the *r^2^* value increased to 0.98 (Figure 4d). Across the top 30 up-or down-regulated peptides that distinguish the COVID-19 patients in Zeno MRM-HR (as in Figure 3c), the average *r^2^* of the mild patient group was 0.53 (up-regulated) or 0.27 (down-regulated) without the standard and improved to an average *r^2^* of 0.88 (up-regulated) or 0.74 (down-regulated) upon OSPP normalization. Also, across all peptides in all severity groups, the median correlation between the normalized quantities obtained with Zeno MRM-HR and ZenoSWATH-DIA improved markedly from 0.39 to 0.83 (Figure 4e). For comparison between methods, we also observed the slope being generally closer to 1 (0.91) when applying normalization to the OSPP internal standard (Figure 4f, Supplementary Table 6). This demonstrate that the use of OSPP in DIA platform helps maintain a robust quantification

Owing to this improvement in analytical precision, the ability to distinguish disease severity also improved and was consistent between the methods. For example, Alpha-2-Macroglobulin (A2M), a broad-spectrum protease inhibitor and an acute-phase protein, was reported to show dropped expression in plasma in COVID-19 in previous studies ^45,46^. In our analysis, the significance of this signal would have been missed in MRM-HR without normalization to the OSPP (*p.adjust* = 0.15 in MRM-HR), but we detected a significant correlation with COVID-19 severity and plasma A2M levels upon normalizing to the OSPP, in both targeted and untargeted proteomics (*p.adjust* = 0.0016 in MRM-HR and *p* = 0.0001 in ZenoSWATH-DIA). In general, 134/199 peptides quantified by MRM-HR and 137/199 peptides quantified by ZenoSWATH-DIA show significant changes with the increase of COVID severity (Figure 4g, Supplementary Table 6).

### Use of the OSPP in comparing plasma proteomic data acquired on different DIA-MS platforms

Since OSPP could work in DIA analysis and support comparable quantification results, we expand the evaluation of OSPP to various LC-MS configurations, spanning from nano-flow chromatography (250 nl/min) to an 800 µl/min analytical flow rate chromatography used in high-throughput applications, coupled to different mass spectrometers, namely an Orbitrap Exploris 480 System (Thermo Scientific) and a ZenoTOF 7600 System (SCIEX). We injected samples from the aforementioned COVID-19 cohort, and data was acquired using DIA-MS on all platforms and processed with the same DIA-NN pipeline.

Out of the 211 peptides of the OSPP, 187 pairs (88.6%) of a native peptide and its matched isotopically labeled internal standard were consistently identified and quantified across more than two-thirds of the samples, across both platforms and different acquisition methods (Figure 5a, Supplementary Table 7). Although the ionisation method and MS acquisition methods are different, we observed a similar distribution of fragments across different platforms (Figure 5 b). To validate the consistency of quantification of each peptide across platforms, we calculated the interclass coefficient correlation (ICC) in samples across platforms. We observed a median ICC value of 0.967 for endogenous peptides and 0.987 for spiked OSPP (Supplementary Table 7), which indicate the consistency of fragment ratio distribution across platforms, showing the robust analytical performance of OSPP peptides in various platforms.

**Figure 5.**
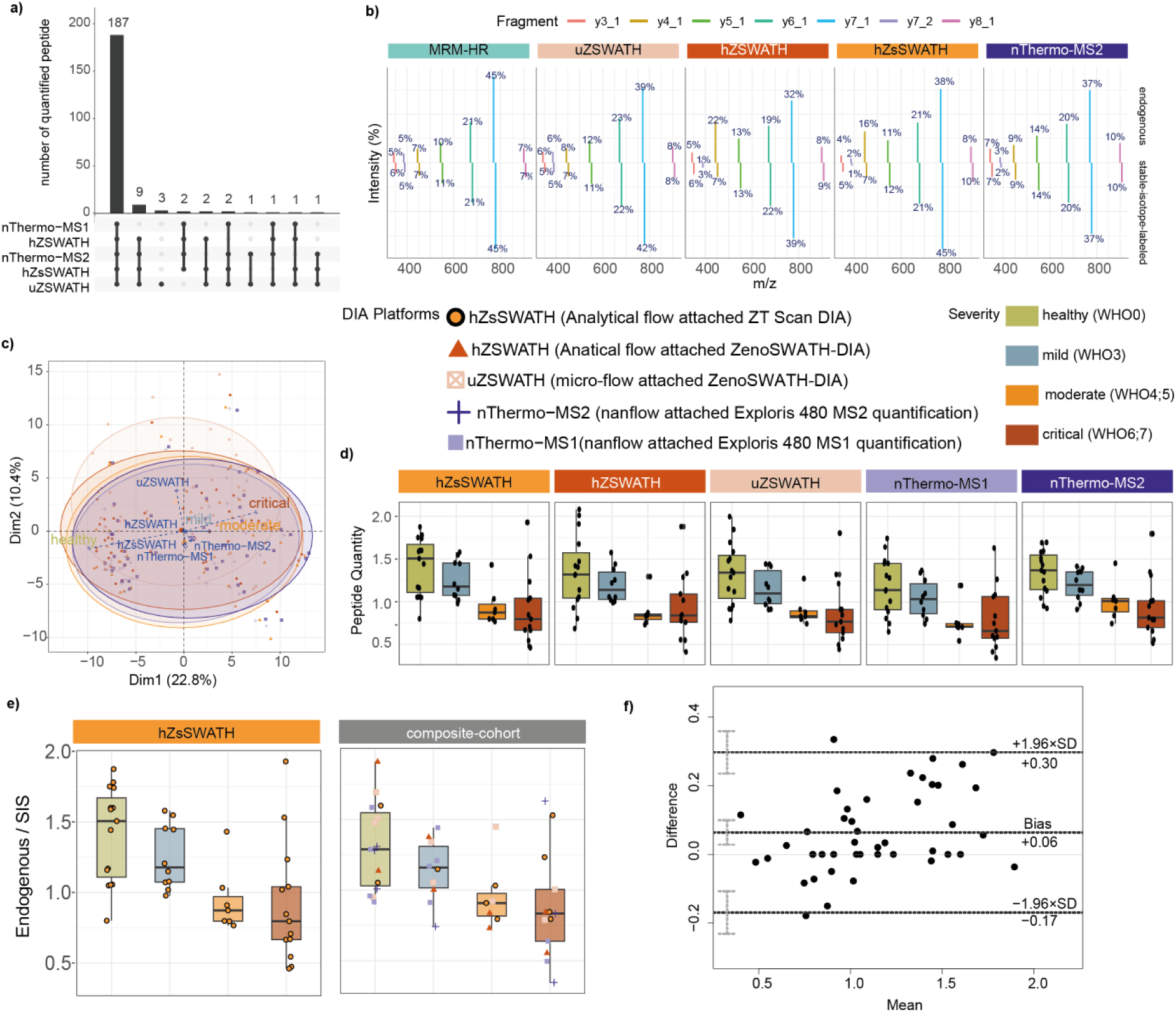
Using the OSPP as an internal standard to align data obtained with the DIA-MS-platform, on the example of a COVID-19 cohort. **a)** OSPP peptides quantified on the indicated DIA-MS platforms (UpSet plot). Intersections of sets of peptides quantified on multiple platforms are shown. Each column corresponds to a DIA-MS platform or set of platforms (dots connected by vertical lines) quantifying the same peptides. The number of peptides in each set appears above the column, while the ones shared per DIA-MS platform are indicated in the graphic below the column, with the name of the DIA-MS platform on the left (nThermo: nanoflow attached Exploris 480, (-MS1: MS1 quantification; -MS2: MS2 quantification), hZSWATH: analytical flow attached ZenoTOF 7600, ZenoSWATH-DIA; hZsSWATH: analytical flow attached ZenoTOF 7600, ZT Scan DIA; uZSWATH: µflow attached ZenoTOF 7600, ZenoSWATH-DIA). **b)** Average fragment spectral distribution of Alpha-2 macroglobulin-derived peptide FEVQVTVPK in triplicate study pool injection in both MRM-HR and ZenoSWATH-DIA. **c)** Unsupervised clustering by principal component analysis (PCA) based on the OSPP normalized quantity of 187 quantified peptides clusters patients with COVID-19 by severity for normalization by the median of endogenous light peptides (left) or via ratio to heavy peptide standard (right). Differences in clustering indicate a reduced influence of technical variance when normalizing via ratio to heavy standard peptide. **d)** OSPP normalized quantities of Alpha-2 macroglobulin-derived peptide FEVQVTVPK across different severity groups. The box-and-whisker plots display 25th, 50th (median), and 75th percentiles in boxes; whiskers display upper/lower limits of data (excluding outliers). **e)** quantities of Alpha-2 macroglobulin-derived peptide FEVQVTVPK in analytical flow attached ZT Scan DIA (hZsSWATH) and “composite cohort” with the same severity distribution in samples from various acquisition platforms. The box-and-whisker plots display 25th, 50th (median), and 75th percentiles in boxes; whiskers display upper/lower limits of data (excluding outliers). **f)** Bland–Altman plot showing the ratio of the mean difference and 95% confidence limits for quantities of all Alpha-2 macroglobulin-derived peptide FEVQVTVPK in analytical flow ZT Scan DIA compared with the “composite cohort” that constitutes samples from various acquisition platforms.

To check the quantification consistency across platforms, we first compared the peptide quantities obtained from study pool samples, which were repeatedly injected at least 3 times on each of the platforms. We noticed that calculating the ratio with the OSPP internal standard value was sufficient to obtain more consistent results across all platforms, with the CV values of all study pool replicates improving significantly for all platforms: without normalization to the OSPP internal standard, cross-platform peptide intensities displayed a median CV of 33.4%, and with normalization to the OSPP, a median CV of 16.2% was observed (Supplementary Table 7).

Next, we conducted a similar analysis with the COVID-19 cohort samples. As expected, peak areas differed between platforms without normalization to the internal standard. Upon forming ratios with the corresponding OSPP standard, the peptides were quantified across platforms with a median CV of just 13.2% (Supplementary Table 7). We also evaluate the consistency of protein quantification across different DIA platforms and distinguish between technical (different acquisition platforms) and biological variability in the dataset using Principal Component Analysis (PCA) (Figure 5c). The analysis revealed that the primary source of variation (PC1, 28.9%) corresponded to the inherent biological differences between samples, as indicated by their separation according to disease severity and treatment escalation scores. In contrast, the secondary source of variation (PC2, 14.7%) was attributed to technical differences between platforms, suggesting that while platform-specific biases exist, they are secondary to the inherited differences of patient samples. The PCA plot confirms that the protein marker quantities effectively classify patients based on treatment escalation scores, underscoring the robustness of the data in reflecting biological differences despite technical variability. Additionally, the loading plot indicates that the key proteins driving the separation along PC1 are those most associated with disease severity, further validating the biological relevance of the analysis. For instance, the peptide FEVQVTVPK derived from broad-spectrum protease inhibitor Alpha-2 macroglobulin is not identified as COVID-19 severity dependent in Zeno MRM-HR without normalizing to OSPP, with the signal intensities varying greatly across DIA platforms. Upon normalization to the OSPP, each of the platforms produces a similar and comparable ratio without technical variance and enables A2M to distinguish COVID-19 severity across platforms (Figure 5d, Supplementary Table 7).

To highlight OSPP’s capability for cross-platform data alignment, we evaluated whether data collected via multiple acquisition platforms can be mixed to produce comparable results to those from a single platform. We created an artificially mixed dataset, a “composite cohort”, reflecting the sample and severity distribution of the original clinical cohort but with peptide quantities randomly selected from various platforms. To make sure if this composite cohort had the same peptide expression changes across severity levels, we matched it with data obtained from an analytical flow combined ZT Scan DIA platform. A total of 89 peptides showed substantial expression changes in both the composite cohort and the single-platform cohort when normalized using the OSPP. Additionally, 20 peptides were significant only in the single-platform cohort, likely due to outliers near the detection limit caused by platform-specific sensitivity differences. Conversely, 6 peptides are significant in other single-platform cohorts also showed significance in the composite cohort rather than the selected single-platform cohort, underscoring the benefits of OSPP normalization. Using OSPP as an internal standard for DIA analysis not only facilitates cross-platform data integration but also improves quantification precision. For example, the peptide FEVQVTVPK from Alpha-2 macroglobulin is significantly upregulated during COVID-19 progression, as observed on the ZT Scan DIA platform (p.adjust = 0.0001), and this trend is similarly confirmed in the composite cohort (p.adjust = 0.0009) (Figure 5e).

To examine he quantification consistency across the two cohorts, we generated Bland-Altman plots, revealing close concordance between the ZT Scan DIA data and the mixed-platform composite cohort data (Figure 5f). Upon normalizing the endogenous peptides using the OSPP, the quantitative data obtained with both cohorts agrees nicely (Bias = 0.06), with few data points falling outside its limits (Figure 5f). The median bias for all peptides across samples is near zero (-0.098), with 96.5% of the peptides showing bias between -1 and 1, signifying significant concordance in peptide ratios between the two cohorts (Supplementary Table 7); Six peptides showing a larger bias when comparing the composite cohort to the single-platform cohort; This discrepancy may stem from the varying sensitivity of the acquisition instruments, which can push certain peptides close to their limits of quantification, thereby generating outliers that impact comparisons. The robust cross-platform consistency underscores OSPP’s potential to provide reliable data comparison and the possibility of data integration across various analytical workflows, hence accentuating its value for mixed-platform data applications and enhancing the overall utility of OSPP in clinical proteomics.

## Discussion

With the increased focus on plasma and serum proteomics, the demand for workflows allowing cross-platform and cross-study comparisons is increasing. While the ultimate goal is to develop analytical technologies that are absolutely quantitative at the protein level, the absolute quantification at the peptide level is easier implement in routine workflows, and substantially enhance analytical precision and accuracy and be used to achieve cross-platform comparability of the generated data ^16,20–22,26,27,33^. In our study, we generate an Open Standard for Plasma Proteomics (OSPP), with the aim of making an internal peptide standard panel that is cost-effective and easily accessible by the community for conducting plasma and serum proteomic experiments in both targeted and untargeted proteomic investigations. Our peptide selection strategy marks a significant advancement in developing robust and versatile panels for quantitative proteomics. While selecting biomarker panels from DIA datasets is a well-established approach—successfully demonstrated in prior studies ^33,47–49^. —our work extends this concept by focusing on a broader scope. Instead of targeting biomarkers for a specific disease, we leveraged large-scale empirical DIA datasets from multiple human cohorts to ensure the selected peptides captured comprehensive information on peptide, and to find peptides that are not disease-specific. Despite these strengths, certain limitations should be acknowledged. For example, all data used for peptide selection were acquired using the same mas spectrometry platform, using SWATH and Scanning SWATH. Albeit we tested the final panel and obtained excellent quantification performance on series of different instruments and acquisition methods, this selection constraint means that peptides that are not well detected on this platform are lacking in our standard

We achieve cost-effectiveness and versatility both by selecting peptides that are consistently detected and quantified with low variance and that have properties that make their synthesis efficient. The 211 peptides included in the OSPP are derived from proteins that function in the blood as part of key biological processes, including metabolism, the innate and adaptive immune system, and the coagulation system, that are changed in a number of diseases (Figure 2b). Indeed, many of these proteins represent established markers available to clinical routine, and several of the included proteins are targets of FDA-approved drugs (Figure 2b). For this reason, the OSPP can be used for targeted proteomics to obtain a broad plasma proteome signature based on the proteins covered. We demonstrate the utility of this approach in the use case of a well-balanced COVID-19 patient cohort, on which we detect biologically meaningful signals within the proteins that are directly covered by the peptides that are represented in the OSPP. For example, the peptide quantities classified the patients according to the disease severity and consequently required treatment according to the WHO ordinal scale, with individual peptides distinguishing different treatment escalations, for example, healthy individuals from mild but hospitalized COVID-19 patients, while other peptides classified moderate from critical patients.

One limitation of any SIS is that a targeted peptide panel does not cover all potential protein biomarkers, nor the protein isoforms of these. However, due to the open nature of the OSPP, it can be customized, for instance, through adding disease- or isoform-specific peptides. In theory, the OSPP can also be mixed with other SIS panels, such as PQ500^23^, which would increase the number of peptides for a broader targeted analysis. Moreover, when applied in discovery proteomic methods, the use of the OSPP or any other SIS does not restrict protein identification to those for which peptides are present in the standard but allows a comprehensive identification of all proteomes while absolutely quantifying the peptides in the standard. Indeed, the use of an internal standard can also be helpful to improve the quantification of peptides that are not covered by the standard, for instance, upon applying cross-normalization strategies. In any case, an internal standard is a helpful tool for batch normalization and quality control and to achieve cross-platform compatibility. Accordingly, simplifying data analysis is an underestimated benefit of SIS panels. In our experience, the hands-on time of bioinformaticians and computational biologists can be a limiting factor in proteome studies. Specifically, the correction for complex batch effects and achieving cross-study comparisons requires specific skills, can be study-specific, and is time-consuming. Herein, we have shown that even a simple strategy of data normalization, like calculating a ratio between endogenous peptides and matching OSPP standards, improves data consistency and mitigates batch effects.

In this study, we used OSPP both by performing calibration series, but also, we tested simple single-point calibration, effectively forming ratios between the endogenous peptides and their standards. Even this simple strategy efficiently minimized technical variance, and could thus be suited for studies that aim for consistent quantification with a minimum time for measurement and data analysis. However, single-point calibration has limitations when larger concentration differences exist or when flexibility in OSPP composition and concentration is needed. In such cases, a calibration using matrix-matched standard dilutions ensured greater accuracy ^50^. While we herein used external calibration curves primarily to estimate limits of quantification (LOQs) for each platform, this approach also allows for a better absolute quantification would improve data comparability.

In conclusion, with the Charité *<u>O</u>pen Peptide <u>S</u>tandard for <u>P</u>lasma <u>P</u>roteomics* (OSPP), we present a highly optimized human SIS panel to be used as an internal standard in plasma and serum proteomic studies. We have shown the standard panel functions in combination with a wide range of analytical platforms and acquisition techniques, including targeted and data-independent acquisition methods on different platforms. On the basis of a COVID-19 inpatient cohort, we validated the OSPP for patient stratification and outcome prediction, using both discovery and targeted proteomics. Including the OSPP enabled consistent quantification across instruments and acquisition methods, allowing composite cohorts from different platforms to achieve clinical readouts comparable to single-platform datasets. Finally, we would like to highlight that the open nature of the standard, the focus on peptides that are easily synthesized and concentration matching, renders the OSPP versatile and cost-effective peptide panel. It can be expanded (i.e. with disease specific peptides) and customized, and thus offers flexibility and precision in high-throughput plasma proteomics with low additional cost per sample. In order to ease the implementation and adoption of OSPP in research applications, we have provided open access to not only the information of peptide standards but also to details on acquisition methods, data processing pipelines, and spectral libraries for DIA proteomics.

## Data Availability

LC-MS acquisition schemes, data analysis pipeline, spectral library as well as dataset used for data analysis and visualization are available online in Mendeley ^51^

## Ethics

The COVID-19 cohort is a subcohort of the Pa-COVID-19 study, which is carried out according to the Declaration of Helsinki and the principles of Good Clinical Practice (ICH 1996) where applicable and was approved by the ethics committee of Charité-Universitätsmedizin Berlin (EA2/066/20).

## Data Availability

All data produced in the present work are contained in the manuscript, or available online at Mendeley Data.

https://doi.org/10.17632/f8kbg4798h.1

## Acknowledgment

We thank all members of Charité Core Facility High Throughput Mass Spectrometry, and Lei Feng (Institute of Forensic Science, Ministry of Public Security, China) for technical support in the preparation of peptides and also Arturas Grauslys (Eliptica) for helping with data analysis. We further thank the Pa-COVID-19 study group^39,40^ for study logistics and collection of biosamples and clinical data. This work was supported by the Ministry of Education and Research (BMBF), as part of the National Research Node ‘Mass Spectrometry in Systems Medicine’ (MSCoreSys), under grant agreements 16LW0239K (to M.M.), 01EP2201 (to M.R.), the Deutsche Forschungsgemeinschaft (DFG, German Research Foundation) – 492697668 and the European Research Council (ERC-SyG-2020 951475). This work was further supported by a BIH Booster Grant (2022-B3020086-11/12 to Z.W., P.T-L., F.K., J.H. M.M, M.R.) and DKTK under grant agreement BE01 1020000483. Z.W. is a member of the International Max Planck Research School (IMPRS) for Infectious Diseases and Immunology, and the Deutsche Forschungsgemeinschaft (DFG) funded Sonderforschungsbereich (SFB) TRR 186. Figure 1 was made using BioRender (Biorender.com).

## Author Contribution

Z.W.:Experimental Design, Data Curation, Sample Preparation, Data Collection, Methodology, Formal Analysis, Visualization, Writing – Original Draft Preparation

V.F.: Formal Analysis, Peptide Selection, Writing – Original Draft Preparation LR.S: Data Collection, Visualization, Writing – Original Draft Preparation

P.T-L.: Sample Collection, Writing – Original Draft Preparation

D.L.: Sample Preparation

F.A.: Data Collection

K.T-T.: Data Collection

A.F.: Data Collection

A.N.: Sample Preparation

AS.W.: Data Collection

AAJ.W.: Data Collection

L.L.: Data collection

F.K.: Consultation, Funding Acquisition

M.S.: Consultation, Supervision

J.H.: Conceptualization, Funding Acquisition, Supervision, Consultation, Writing – Original Draft Preparation

M.M.: Conceptualization, Funding Acquisition, Project Administration, Resources, Supervision, Consultation, Writing – Original Draft Preparation

M.R.: Conceptualization, Funding Acquisition, Project Administration, Resources, Supervision, Consultation, Writing – Original Draft Preparation

## Materials and Methods

### Reagents

Water was from Merck (LiChrosolv LC-MS grade; Cat# 115333), acetonitrile from Biosolve (LC-MS grade; Cat# 012078), trypsin (Sequence grade; Cat# V511X) from Promega, 1,4-Dithiothreitol (DTT; Cat#6908.2) from Carl-Roth, iodoacetamide (IAA; Bioultra; Cat# I1149) and urea (puriss. P.a., reag. Ph. Eur.; Cat#33247) were from Sigma-Aldrich, ammonium bicarbonate (Eluent additive for LC-MS; Cat# 40867) and Dimethyl sulfoxide (DMSO; Cat# 41648) were from Fluka, formic acid (LC-MS Grade; Eluent additive for LC-MS; Cat# 85178) was from Thermo Scientific™, bovine serum albumin (BSA; Albumin Bovine Fraction V, Very Low Endotoxin, Fatty Acid-free; Cat# 47299) was from Serva., commercial human plasma samples (Human Source Plasma, LOT# 20CILP1034) was from zenbio.

### Peptide Selection and Synthesis

To prioritize the most reliably quantified precursors and minimize the influence of such factors as precursor abundance, study cohort, MS setups, LC separations, and sample preparation procedures, we introduced a relative rank metric, which was defined as following. First, we defined precursor weight as a ratio of a precursor’s % presence *PPres*, to the coefficient of variation %*CV*

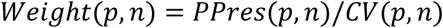

and a weight-based rank *Rank*(*p*, *n*) = *rank*{*Weight*(*p*, *n*)}. Here, *p* stands for precursor and *n* for a study pool series. The weight thus corresponds to a precursor’s signal-to-noise ratio (*S*/*N* = 1/*CV*) multiplied by its presence. To minimize the influence of the total number of precursors on the ranking, we introduced relative rank *RelRank*(*p*, *n*), defined as the ratio of the precursors rank to the maximum rank value in a study

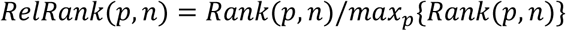

Finally, the precursor’s average (over considered studies) relative rank *RelRank*(*p*) was used to select the best „global“ (i.e. non-project specific) precursors for every protein while we also required that the lower cutoff of the relative rank be set as 0.6.

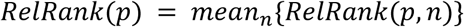

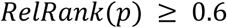

Additionally, we only consider proteotypic peptides in our panel and for more reliable quantification require those peptides quantified in at least half of the projects:

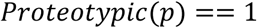

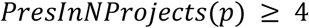

To avoid all peptides coming from those top abundant proteins in plasma and to allow covering a larger dynamic concentration range of proteins, only the top 3 peptides are selected for each protein.

#### Further selection based on physical-chemical-and analytical properties

The chemical properties of each peptide are calculated by the R package “Peptides v2.4.6”. The hydrophobicity of each peptide is calculated by function “hydrophobicity_kyte” ^52^, the hydrophobicity scales run from -2 to 2 where 94 peptides are hydrophobic (>0) and 117 are hydrophilic (<0); net charge is calculated with function “charge”; high missed cleavage is considered and excluded when “KK|KR|RR|RK|KP|RP” appears in the peptide sequence, with the exception of peptide “AN<u>R</u>PFLVFIR” (SERPINC1) which we previously found to be of interest and with good performance across large numbers of samples ^33^. Peptides containing cysteine and N terminal glutamine that are easily modified are excluded except “I<u>C(Carboxymethylated)</u>LDLQAPLYK” which is the only selected peptide for protein “PF4”. An additional 24 peptides (30 peptides, 6 of which are also selected from previously mentioned study pools selection) from the previous MRM panel ^33^ were included in the list. For checking the synthesis possibility of peptides, the Peptide Synthesis and Proteotypic Peptide Analyzing software tool (Thermo, [34]) was used with synthesis.

A pool of all the study pools used for the initial selection was prepared and analyzed on a 20-min water-to-acetonitrile 5µl/min microflow-rate chromatographic gradient analyzed by high-resolution multiple reaction monitoring (Zeno MRM-HR) on a ZenoTOF 7600 instrument (SCIEX) to check the analytical performance of all shortlisted peptides. Nearly all peptides were well identified with a charge state mostly 2 or 3. One peptide EGPYSISVLYGDEEVPRSPFK from protein FLNA failed to be identified on µflow, however, it has a good identification on the analytical flow LC attached MS instrument with a charge state of 4.

All the above criteria are listed in Supplementary Table 1.

#### Peptide Synthesis and validation

Reference peptide standards were custom synthesized by Pepmic Co., Ltd (Suzhou, China) where native peptides (natural, light [NAT]) were obtained at ≥95% purity and stable isotope-labeled heavy labeled peptides (labeled on C-terminal lysine (K) or arginine (R) with stable isotopes (K(U-^13^C_6_,^15^N_2_) or R(U-^13^C_6_,^15^N_4_))) - at ≥70% purity. Validation of the synthesized peptides involved initial assessment via LC-UV/VIS and LC-MS analysis.

All peptide stock solutions were prepared at 1 mg/ml in 50:50 (v/v) ddH2O: acetonitrile mix. The peptides were batch-pooled in groups of 11 (∼20 peptides per group) of each native and isotopic labeled standard, based on their endogenous abundance in the EDTA plasma pool of all the study pools acquired by µ-flow DIA MS. The peptide pools were further analyzed on the same LC-MS method. The validation of peptide synthesis is considered in two aspects: all isotopically labeled peptides should coelute with their corresponding native forms in chromatograms, and no native peptide was identified in isotopically-labeled-only pools, confirming the satisfactory purity of approximately 70% and affirming their successful synthesis and compatibility with our analytical platform. All synthesis peptide standards passed the above criteria and are aliquoted and stored in 96-well plates in a -80°C freezer for future preparation.

### Generation of the OSPP mixture

We first mixed all isotopically labeled heavy peptide standards to reach a final concentration of 1 µg/µl and conducted dilution series w. 1/10/100/300/900 pg/µl of each peptide. The signal ratio (native endogenous peptide signal / heavy isotope labeled peptide signal) is calculated for each peptide in each concentration. For selecting an appropriate concentration of each peptide, we first calculate the linearity range of each peptide within 1-900 pg/µl of concentration. Among the linear concentrations, only the concentrations where heavy peptide quantities closely match their native counterparts within a 2x log10 difference were chosen. The concentration of each peptide was further adjusted and calculated to make sure all heavy peptides’ signals were the same or at most within a log10 difference from their endogenous counterparts. Next, we categorized all peptides into four distinct concentration tiers, mixing to establish a comprehensive concentration range of 10 pg/µl to 2 ng/µl of each peptide within the OSPP mixture(Supplementary Table 2). To avoid possible evaporation, the OSPP are dilated in 10% w/v acetonitrile, exhibiting no discernible evaporation effects when mixed with digested plasma samples in 384-well plates. We also tested the performance of the OSPP by spiking 1 µl (40.4 ng for all 211 peptides) into every 1.5 µg of digested plasma pool; signals of all peptides fell within log10 difference to their respective endogenous signals.

#### Equally-concentrated (“Single-conc. Std”)

“Single-conc. Std” was prepared by pooling the same amount of each peptide. In the mixture, all peptides are equally concentrated with 600 pg/µl of each, and housed in 50% Acetonitrile. For matrix performance tests, the single-conc. Std was in 100 pg/µl as diluted in 10% Acetonitrile.

### Sample Preparation

#### Plasma Samples & BSA

Samples were prepared with minor modifications as described previously ^44^. Briefly, plasma/serum samples were stored at -80°C for 11-12 months prior to preparation, and clinical samples and calibration series were prepared as follows: 5 µl of citrate plasma were added to 55 µl of denaturation buffer, composed of 50 µl 8 M Urea, 100 mM ammonium bicarbonate, 5 µl 50 mM dithiothreitol (DTT) and internal standard mix. The samples were incubated for 1 h at room temperature (RT) before the addition of 5 µl of 100 mM iodoacetamide (IAA). After a 30 min incubation at RT, the samples were diluted with 340 µl of 100 mM ammonium bicarbonate and digested overnight with 22.5 µl of 0.1 µg/µl trypsin (ca. 1:150 (m/m) Trypsin: Substrate ratio) at 37°C. The digestion was quenched by adding 50 µl of 10% v/v formic acid. The resulting tryptic peptides were purified on a 96-well C18-based solid phase extraction (SPE) plate (BioPureSPE Macro 96-well, 100 mg PROTO C18, The Nest Group). The purified samples were resuspended in 120 µl of 0.1% formic acid. 1 µl of OSPP was spiked to 1.5 µg of digested plasma and injected on LC-MS/MS platforms (ZenoTOF 7600, timsTOF, Exploris480) at customized volumes.

#### Calibration Curves

We introduce an 8-point calibration curve with BSA as a surrogate matrix. For the seven non-zero calibration samples, 10 µl of the OSPP mixture (same as what the sample are used) was mixed with 10 µl of a dilution series of the native peptide standard pool ranging from 1000 to 0.064 pg/µl; 20 µl of BSA tryptic digest was then added as a surrogate matrix. The last sample of the calibration series used 10% (v/v) acetonitrile buffer instead of the light peptide standard (see details in Supplementary Table 7).

### Liquid chromatography Mass spectrometry

#### Micro-flow-rate (µflow) LC attached ZenoTOF 7600 (ZenoSWATH-DIA, Zeno MRM-HR)

All samples were acquired on an ACQUITY UPLC M-Class system (Waters) coupled to a ZenoTOF 7600 mass spectrometer with an Optiflow source (SCIEX). Prior to MS analysis, 250 ng samples were loaded onto LC and chromatographically separated with a 20 min gradient (time, % of mobile phase B: 0 min, 3%; 0.86 min, 7.1%; 2.42 min, 11.2%; 5.53 min 15.3%; 9.38 min, 19.4%; 13.02 min, 23.6%; 15.48 min, 27.7%;17.27 min, 31.8%; 19 min, 40%; 20 min, 80% followed by re-equilibration for 10 min before the next injection) on a HSS T3 column (300 µm×150 mm, 1.8 µm, Waters) heated to 35°C, using a flow rate of 5 µl/min where mobile phases A and B are 0.1% formic acid in water and 0.1% formic acid in acetonitrile, respectively. To avoid introducing technical variance due to differences in injection volumes, we always injected a constant volume of the plasma sample or calibration series samples.

### ZenoSWATH-DIA

A Zeno SWATH acquisition scheme with 85 variable-sized windows and 11 ms MS2 accumulation time was used. Ion source gases 1 and 2 were set to 12 and 60 psi, respectively. Curtain gas was at 25 psi, CAD gas at 7 psi, and source temperature was set to 300°C; spray voltage was set to 4500 V.

### Multiple Reaction Monitoring - High resolution (Zeno MRM-HR)

A scheduled Zeno MRM-HR method with identical instrument setting parameters as for Zeno SWATH was developed and used. The choice of precursor and selection of retention time was adopted based on triplicate injections of EDTA plasma sample on the microflow attached ZenoSWATH-DIA. The Zeno threshold was set to 20,000 cps and for all peptides, the TOF MS2-scan range was from 200 to 1500 m/z, respectively. MS2 accumulation time was set to 13 ms. Retention time tolerance was set as +/- 20 seconds. Collision energies were defined based on the following formula: CE = slope * m/z + intercept, Supplementary Table 8).

**Supplementary Table 8.** calculation formula for Zeno MRM-HR method.

| Charge State | Slope | Intercept |
| --- | --- | --- |
| 1 | 0.05 | 5 |
| 2 | 0.049 | -1 |
| 3 | 0.048 | -2 |
| 4 | 0.05 | -2 |

#### Analytical flow-rate system LC attached ZenoTOF 7600 (ZenoSWATH-DIA MS, ZT Scan DIA)

Samples were acquired on a 1290 Infinity II UHPLC system (Agilent) coupled to a ZenoTOF 7600 mass spectrometer with a DuoSpray TurboV source (SCIEX). Prior to MS analysis, samples were chromatographically separated on an Agilent InfinityLab Poroshell 120 EC-C18 1.9 µm, 2.1 mm × 50 mm column heated to 50°C. A gradient was applied that ramps from 3 to 36% buffer B in 3 min (buffer A: 1% acetonitrile and 0.1% formic acid; buffer B: acetonitrile and 0.1% formic acid) with a flow rate of 800 µl /min. For washing the column, the flow rate was increased to 1.2 ml /min and the organic solvent was increased to 80% buffer B in 0.1 min and was maintained for 1.4 min at this composition before reverting to 3% buffer B in 0.1 min. 1.5 µg of the plasma sample or calibration series sample was loaded prior to cohort samples entering MS.

**ZenoSWATH-DIA** acquisition scheme with 60 variable-sized windows and 13 ms MS2 accumulation time was used. Ion source gas 1 (nebulizer gas), ion source gas 2 (heater gas), and curtain gas were set to 60, 65, and 55 psi, respectively; CAD gas was set to 7 psi, source temperature to 600°C, and spray voltage to 4000 V.

The **ZT Scan DIA** method used the same instrumental source setup parameters as ZenoSWATH-DIA. The method consisted of an MS1 scan from m/z 100 to m/z 1000 and 25 MS2 scans (25 ms accumulation time) with variable precursor isolation width covering the mass range from m/z 400 to m/z 910. Q1 mass width is set as 2.5 Da with a scan speed of 750 Da/Sec. The applied collision energies were as for Zeno SWATH (derived from a linear equation, see above).

#### Analytical flow-rate system LC attached timsTOF HT

Samples were analyzed on a Bruker timsTOF HT mass spectrometer coupled to a 1290 Infinity II LC system (Agilent). Before MS detection, 5 µg of the sample were chromatographically separated on a Phenomenex Luna®Omega column (1.6 μm C18 100A, ^53^ 30 × 2.1 mm) heated to 50°C, using a flow rate of 0.5 ml/min where mobile phase A & B were 0.1% formic acid in water and 0.1% formic acid in acetonitrile, respectively. The LC gradient ran as follows: 1% to 36% B in 5 min, increase to 80% B at 0.8 mL over 0.5 min, which was maintained for 0.2 min and followed by equilibration with starting conditions for 2 min.

For **diaPASEF MS** acquisition, the electrospray source (Bruker VIP-HESI, Bruker Daltonics) was operated at 3000 V of capillary voltage, 10.0 l/min of drying gas, and 240 °C drying temperature. The diaPASEF windows scheme was as follows: we sampled an ion mobility range from 1/K0 = 1.30 to 0.7 Vs/cm2 using ion accumulation times of 100ms and ramp times of 133ms in the dual TIMS analyzer, each cycle times of 1.25 s. The collision energy was lowered as a function of increasing ion mobility from 59 eV at 1/K0 = 1.6 Vs/cm2 to 20 eV at 1/K0 = 0.6 Vs/cm2. For all experiments, TIMS elution voltages were calibrated linearly to obtain the reduced ion mobility coefficients (1/K0) using three Agilent ESI-L Tuning Mix ions (m/z, 1/K0: 622.0289, 0.9848 Vs/cm2; 922.0097, 1.1895 Vs/cm2; and 1221.9906, 1.3820 Vs/cm2).

#### Nanoflow rate LC attached Exploris 480 (Thermo Scientific)

Samples were analyzed on an Exploris 480 (Thermo Scientific) coupled to a Vanquish Neo UHPLC-System (Thermo Scientific) utilizing a 22-minute gradient in nanoflow (0.25µl/min). For LC separation, the attached column was an in-house packed 20 cm long 1.9 µm column. A shortened gradient time was used with the published acquisition method ^54^ where mobile phases A & B were 0.1% formic acid plus 3% acetonitrile in water and 0.1% formic acid in 90% acetonitrile, respectively. The LC gradient ran as follows: increased from 2% buffer B to 30% buffer B over the course of the first 14.5 minutes and increased to 60% buffer B within the next 1.5 minutes. Finally, buffer B concentration increased to 90% for one minute and was held for 5 minutes to flush the column.

For Orbitrap acquisition, full scans were acquired between 350-1650 m/z with a resolution of 120,000. For MS2 scans, the maximum injection time was set to 54 ms, and scans were made over 40 variable-sized isolation windows.

### Generation of OSPP-specific Human Spectral Library

A comprehensive spectral library for human Stable Isotope Labeling was constructed through a multistep process using DIA-NN and a custom R script. For all the experiments, we used a project-independent public spectral library DiOGenes ^31^ reannotated by Human UniProt ^55^ (UniProt Consortium, 2019) isoform sequence database (3AUP000005640, [27 March 2023]). The library was first automatically refined based on the dataset at 0.01 global q-value (using the “Generate spectral library” option in DIA-NN). DIA-NN was employed with specific commands to enhance the library’s accuracy and utility and label all Arginines and Lysines in the existing spectral library: --fixed-mod SILAC,0.0,KR, label --lib-fixed-mod SILAC --channels SILAC,L,KR,0:0; SILAC,H,KR,8.014199:10.008269 --peak-translation --original-mods -- matrix-ch-qvalue 0.01

This set of commands facilitated the automatic segregation of the spectral library into multiple channels, particularly for precursors associated with the Lysine and Arginine label group modification. To improve precision and accuracy during quantification, this heavily labeled spectral library was further refined. This refinement involved only keeping the label for peptides from OSPP with only the C terminal Lysine or Arginine labeled; and for quantification accuracy, all b-ions were excluded from quantification by labeling b-ions as “T” in the “ExcludeFromAssay” category.

### Data Acquisition & Processing

All raw data from the ZenoTOF 7600 system were acquired by SCIEX OS (v. 3.0). All raw data from timsTOF HT were acquired with timsControl (v.5.1.8) and HyStar (v.6.3.1.8). All raw data from Exploris 480 (Thermo) were acquired using Xcalibur.

#### Discovery proteomics

The raw proteomics data from all DIA methods was processed using DIA-NN, 1.8.1, available on GitHub (DIA-NN GitHub repository ^56^). The MS2 and MS1 mass accuracies were set to 20 and 12 ppm (ZenoTOF 7600 data) or 15 and 15 ppm (timsTOF and Exploris 480 data), and the scan window to 7. The aforementioned OSPP-specific Human Spectral Library is used for data processing with additional commands: --fixed-mod SILAC,0.0,KR,label --channels SILAC,L,KR,0:0; SILAC,H,KR,8.014199:10.008269 --peak-translation --original-mods -- matrix-ch-qvalue 0.01 --restrict-fr --report-lib-info

Specifically, following a two-step MBR approach ^30^, an in silico spectral library is first generated by DIA-NN from the FASTA file(s); this library is then refined based on the DIA dataset and subsequently used to reanalyze the dataset, to obtain the final results.

The data were filtered in the following way. First, a 1% run-specific q-value filter per isotope channel was automatically applied at the precursor level by DIA-NN (--matrix-ch-qvalue 0.01). We note that in any experiment processed using the MBR mode in DIA-NN, 1% global precursor q-value filtering is also applied automatically ^30^.

For quantification, we used “Precursor.Translated” value as quantities for each precursor in MS2 quantification. For Exploris 480 data, since orbitraps are sensitive in MS1, we also used “Ms1.Translated” was used.

#### Targeted proteomics

Zeno MRM-HR data were processed using Skyline (64-bit, v.23.1.0.268). No blinding was performed during peak integration. The quantity of each peptide is calculated by the summation of peak areas of each selected fragment of a peptide (list of fragments used for quantification in Supplement Table 4).

#### Calibration curve

The calibration curve for each of the 211 peptides was either accepted or rejected based on a set of rules and criteria: the limit of quantification (-LLOQ and –ULOQ) was determined based on the accuracy of replicated injection on the same LC-MS platform (Supplementary Table 6). Peptide concentration (expressed in pg/µl) was determined from calibration curves, constructed with native and isotopic labeled peptide standards in the surrogate matrix (4 ng/µl BSA), and manually inspected and validated. Peptides with > 40% of values below the lowest or above the highest detected calibrant concentration across all samples were removed from the analysis. Linear regression analysis of each calibration curve was performed using custom R code (with 1/x weighting).

### Data Analysis

#### Data Completeness

The completeness of data for each peptide was evaluated based on its frequency of detection across all biological samples. Peptides were considered if they were detected in more than 66.7% (⅔) of the samples. We calculated the percentage of each peptide measured on each LC-MS platform/method and used only the peptides with a completeness value exceeding 66.7% for subsequent analysis.

#### Data Normalisation

Two normalizing strategies to evaluate the quantification consistency were applied. The first approach was a normalization by median division of all endogenous peptide quantities in the study pools (except in the Thermo instrument, replicate 04 is excluded due to acquisition failure) measured on each platform. All peptides in each platform were applied with this factor, referred to as “norm_light”.

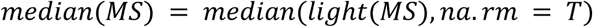

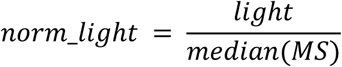

In addition, with the spiked OSPP mixture, we use the heavy isotope-labeled spiked peptide standard in each sample to normalize the corresponding endogenous peptide levels in the sample (endogenous peptide quantitym (light) /quantity of correspondent heavy labeled peptide quantity (SIS)), termed as “ratio”.

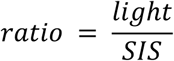

#### Precursor Selection

As several precursors (charge state of +1 to +4) from the same peptide are quantified on different platforms, several criteria should be fulfilled to choose the best precursor used for follow-up quantification and cross-method / cross-platform comparison: a) Due to the difference in analyte ionization ability on various MS platforms, different precursors from the same peptide will show various abundances, the most abundant one shall be the charge state with the best ionization efficiency. b) Moreover, the abundance will also affect the reproducibility of the performance of isotopically labeled peptide standards. For replicate injections of study pool samples on each platform, we filtered for precursors with CV less than 40% to guarantee reproducibility. c) Additionally, for precursors of different peptides from the same protein, we checked the behavior of isotopic labeled peptide standards throughout all study samples and only chose the precursor that showed the same trend. The precursors used for quantification on different MS platforms are listed in Supplementary Table 6.

A coefficient of variation (CV) was calculated for each precursor as its median absolute deviation (R “stats v4.2.2” - function “mad()”) divided by its empirical median and multiplied by 100 to report in percentages.

#### Statistical analysis and visualization

Significance testing of the trend between absolute peptide concentrations and the ordinal classification as provided by the WHO disease severity(levels as indicated) was performed using Kendall’s tau (KT) statistics as implemented in the “EnvStats v2.8.1” R package “kendallTrendTest” function. For cohort 2 the KT statistics were calculated as the trend of absolute peptide concentrations against the following WHO groups: 0, 3, 4, 5, 6, and 7; selected peptides in each comparison were used for data analysis, without imputation. Where indicated, multiple testing correction was performed by controlling for false discovery rate using the Benjamini-Hochberg procedure 1 as provided by the R package “stats v4.2.2” - “p.adjust” function. A full summary of these statistical test results is provided in respective Supplementary Tables. (Adjusted) P values were considered significant when *p* < 0.05.

The upset plot is visualized using “UpSetR v.1.4.0” - function “upset”; the Venn diagram is visualized using “ggvenn v0.1.10”. Principal component analysis was performed and visualized using the R function “fviz_pca_biplot” from package “factoextra v1.0.7”. All other visualization is performed using “ggplot2 v.3.4.4”

